# Exploring Digital Health Solutions: Personalised Medicine and N-of-1 Trials in Ghana: A Scoping Review

**DOI:** 10.1101/2024.12.29.24319752

**Authors:** Godsway Sackey, Babajide Owoyele, Frank Baiden, Stefan Konigorski

## Abstract

**Background:** Digital health solutions and personalised medicine are rapidly evolving fields that offer significant potential for enhancing healthcare delivery, particularly in low-resource settings. N-of-1 trials, a personalised experimental approach, hold particular promise for individualised patient care and empowerment. This scoping review aims to explore the current landscape of these innovations in the context of Ghana.

**Objectives:** This review seeks to map existing literature on digital health solutions, personalised medicine, and N-of-1 trials in Ghana. It also aims to identify key themes, trends, and gaps in the literature while discussing the current role and potential of N-of-1 trials in this context as an important knowledge base for future exploration of personalised medicine and digital N-of-1 trials in low resource settings like Ghana.

**Methods:** A comprehensive literature search was conducted across multiple databases, including PubMed, Scopus, and Google Scholar; from year 2000 to April 2024, to identify relevant studies. Inclusion criteria were studies focusing on digital health, personalised medicine, and N-of-1 trials within Ghana or similar low-resource settings. Data were extracted and analysed to identify common themes, trends, and gaps in the existing literature.

**Results:** The scoping review included 40 publications, that is, only very few studies have been published in this field that fit our objectives. The studies revealed a predominant focus on digital health solutions in Ghana, particularly mobile health (mHealth) interventions, which account for 60% of the interventions reviewed. These solutions include mobile applications for clinical decision support, patient monitoring, and health education, and have been effective in enhancing healthcare delivery, especially in remote and underserved areas. Introducing electronic health records (EHRs) represent 24% of the interventions and have shown promise in improving data management and patient care coordination. However, challenges such as poor network infrastructure and resistance to change need to be addressed. Personalised medicine, though less represented in the reviewed literature (7.5%), shows promise in providing tailored treatments based on individual genetic profiles. The review did not identify any studies specifically focused on N-of-1 trials, highlighting the innovative potential for future research in this area.

**Conclusion:** The findings underscore the need for robust digital infrastructure, training for healthcare providers, and policy frameworks to support the adoption of digital health solutions, personalised medicine, and N-of-1 trials in Ghana. Addressing these gaps requires concerted efforts to enhance digital infrastructure, provider education, and supportive policies. N-of-1 trials offer substantial potential for personalised care but necessitate further exploration and integration into the healthcare system.

## Introduction

Digital health solutions encompass a variety of technologies aimed at enhancing healthcare delivery and outcomes through digital tools, including mobile health (mHealth), health information technology (IT), wearable devices, telehealth, and telemedicine. These technologies can facilitate real-time data collection, remote patient monitoring, and improved diagnostic accuracy. The primary goals of digital health are to increase access to healthcare, reduce costs, and provide personalised care through computing platforms, connectivity, software, and sensors (1–3).

Precision medicine aims to provide tailored treatments based on individual genetic, environmental, and lifestyle factors, leading to more effective and precise medical interventions (4,5). It is sometimes used as an interchangeable term to personalized medicine, but in general refers to a specific subfield by assuming that stratifying results from large population studies by refined group characteristics can yield personalized treatment strategies. Both approaches aim to provide more effective and targeted therapies, minimise adverse effects, and enhance patient outcomes. Key technologies include genomic sequencing, biomarker analysis, and advanced data analytics (6,7). N-of-1 trials are one specific approach for personalized medicine focusing on determining optimal treatment for an individual through repeated crossover trials of different treatments, making them suitable for resource-limited settings like Ghana (8,9). Single patients serve as the entire study population and can be closely monitored in the trials to determine the most effective customised therapy (10,11).

### Historical Developments

The field of digital health has significantly evolved over the past decades, driven by technological advancements and increasing integration of digital tools in healthcare. Key milestones include the rise of telemedicine, the use of mobile health (mHealth) applications, and the development of electronic health records (EHRs). These technologies have enhanced patient care, improved health outcomes, and increased accessibility to healthcare services. Notable developments include early telemedicine technologies over 100 years ago, the integration of wearable devices for health monitoring in the 1940s, and the complete digitization of health data recommended by the US National Academy of Medicine in the mid-1990s (10–13).

The concept of personalised medicine and of precision medicine dates to ancient times, with Hippocrates emphasising individualised treatment. Major milestones include the Human Genome Project (1984–2002), which mapped the entire human genome, and the introduction of targeted therapies and companion diagnostics.

N-of-1 trials have emerged as a valuable tool in personalised medicine, allowing for the evaluation of treatment effects in individual patients through multiple crossover comparisons. The development of these trials has been driven by the need for personalised treatment strategies and the limitations of traditional clinical trials. Key developments include the use of N-of-1 trials in various medical conditions and their potential for fast-tracking drug development in precision oncology (14).

### Current Trends and Advancements

Digital health solutions have seen significant advancements globally, driven by innovations in mobile health (mHealth), big data analytics, and artificial intelligence. These technologies are enhancing healthcare delivery through real-time monitoring, personalised medicine, and improved data management. Key trends include the integration of wearable devices for health monitoring, the rise of telemedicine for remote diagnostics, and the use of big data for predictive analytics in public health (15–17).

In low-resource settings, digital health solutions focus on improving healthcare accessibility and quality. Mobile health technologies, such as SMS-based health information and mobile apps for disease monitoring, have shown significant promise. These technologies help bridge healthcare delivery gaps by providing remote diagnostics, health education, and disease surveillance. Key advancements include scalable digital health programs tailored to local needs, effective integration of digital tools with existing health systems, and the use of digital platforms for health worker training and support (18–20).

### Role and Significance of N-of-1 Trials

N-of-1 trials can play a crucial role in personalised medicine by allowing for a direct evaluation of treatment effects on an individual patient. These trials involve repeated crossover comparisons of different treatments within the same patient, providing highly specific data on how a particular patient responds to each treatment. This approach is especially useful where large-scale randomised controlled trials (RCTs) are impractical, such as with rare diseases or significant individual variability in treatment response (21,22).

N-of-1 trials can provide robust evidence for tailoring treatments to individual patients, improving the precision and effectiveness of medical interventions. They can help identify the best therapeutic option for a specific patient, minimise the risk of adverse effects, and enhance treatment outcomes (23). These trials assist clinicians in making evidence-based decisions tailored to the patient’s unique response, thus integrating personalised care into routine clinical practice. They are particularly beneficial in studying rare diseases where traditional RCTs are not feasible due to small patient populations (24).

### Relevance to Ghana and Low-Resource Settings

Ghana has made significant strides in digitalising its health sector, including implementing national e-health strategies and various pilot projects aimed at improving healthcare delivery. These initiatives include using mobile technologies for data collection, education, and telemedicine services. However, challenges such as a lack of skills, human resources, and technology infrastructure issues remain significant barriers (16,25). The Mobile Technology for Health (MOTECH) program in Ghana exemplifies the application of mobile technologies to improve maternal and child health, despite technical challenges like message delivery and user engagement (26).

The deployment of drone technology for emergency medical deliveries has significantly impacted healthcare delivery, particularly in remote areas. This initiative, part of the "Ghana Go Digital Agenda," has improved the accessibility and timeliness of medical supplies (27). The development and use of digital platforms for national health insurance services have been influenced by various institutional factors. These platforms aim to enhance the efficiency and accessibility of health insurance services (28).

However, poor internet connectivity and erratic power supply are major impediments to the effective implementation of digital health technologies in Ghana. These issues cause delays and disruptions in healthcare delivery, undermining the potential benefits of digital health initiatives (29). There is a need for continuous digital education and training for healthcare providers to enhance their capacity to utilise digital health technologies effectively. Addressing gaps in digital skills and promoting stakeholder engagement are crucial for the success of digital health initiatives (30).

Personalised medicine faces significant challenges in low-income countries, including the high cost of diagnostic techniques and targeted treatments. Integrating traditional medicine with biomedical health systems can expand the reach and improve outcomes of community healthcare by leveraging the extensive infrastructure of traditional medicine (31). Integrating personalized medicine within Ghana’s healthcare system does encounter significant socioeconomic challenges as other similar environments, including high diagnostic costs and access disparities. Alemayehu et al. (3) emphasize that patients in low-income settings face considerable financial burdens, limiting scalability. Solutions to these barriers could include subsidies, partnerships, and localized diagnostic technologies that make personalized care more accessible and feasible (3,4). Personalised medicine plays a crucial role in cancer treatment by allowing for the identification of genetic predispositions and the development of molecularly targeted therapies. These approaches can be adapted to low-resource settings with appropriate support and infrastructure (31). Alemayehu et al. (3) review examples of N-of-1 trials conducted in low-resource settings like China and Brazil. These examples demonstrate the effectiveness of N-of-1 trials in improving patient outcomes affordably. With adequate policy support and regional partnerships, Ghana could adopt similar methods, enhancing patient outcomes while addressing local challenges sustainably.

### Purpose of the Scoping Review

This scoping review aims to explore digital health solutions, personalised medicine, and N-of-1 trials in Ghana, a low-resource setting, to understand their potential prospects and how to integrate these innovations into the current health systems. The review will also serve as a precursor and provide baseline information for subsequent studies in these areas and consider how these advancements can be extended to other low-resource settings to improve healthcare delivery and outcomes.

## Methods

### Study Design and Framework

This scoping review follows the PRISMA-ScR checklist and explanation by Tricco et al. (2018), ensuring a comprehensive and systematic approach (Supplementary Table 1). The scoping review was pre-registered and its protocol is available at https://osf.io/n2krw. Supplementary Figure 1 gives an overview of the review and its components.

### Eligibility Criteria

#### Inclusion Criteria

This scoping review focused on studies examining digital health solutions and personalized medicine, with a specific interest in research addressing the role and implementation of N-of-1 trials within healthcare. To be included, studies had to provide meaningful insights into healthcare practices within low-resource settings, particularly in Africa or comparable environments. We included empirical studies, systematic reviews, and case studies that illuminated the challenges, barriers, facilitators, and successes of implementing digital health and personalized medicine innovations. Eligible studies were required to be available in English, published from the year 2000 onward, and sourced from reputable databases such as PubMed, Scopus, Web of Science, IEEE Xplore, AJOL, the Cochrane Library, Google Scholar, and relevant government or organizational websites. The review prioritized research offering actionable insights, strategic frameworks, or technological innovations relevant to healthcare systems like Ghana’s.

#### Exclusion Criteria

Studies were excluded if they did not specifically focus on digital health solutions, personalized medicine, or N-of-1 trials, or if they lacked relevance to low-resource contexts. We excluded literature that failed to provide empirical data or evidence-based insights for healthcare improvement, as well as publications predating the year 2000. Articles in languages other than English, grey literature, editorials, opinion pieces, and non-peer-reviewed materials were also outside the scope of this review. Additionally, studies from high-resource settings that lacked transferrable lessons or actionable strategies for the focus of this review were not included.

### Search Strategy

The search strategy was developed by our team to ensure comprehensive coverage, see Supplementary Text 1 for details. The databases searched included PubMed, Directory of Open Access Journals, IEEE Xplore, the Cochrane Library, Google Scholar, and relevant government and company websites from 2000 until April 2024. Keywords and search terms were derived from the main themes of the review including digital health, personalized medicine, and N-of-1 trials tailored to the context of Ghana and low-resource settings.

### Study Selection

The selection process involved two stages: initial screening of titles and abstracts followed by a full-text review. We reviewed included studies independently and screened them for inclusion. We resolved discrepancies and followed PRISMA flow diagram to illustrate the entire study selection process.

#### Stage 1: Initial Screening of Titles and Abstracts

We independently screened the titles and abstracts of all retrieved studies. This was done purposely to identify studies that potentially met the inclusion criteria. For example, studies such as "Using Mobile Health to Support Clinical Decision-Making in Ghana" (32) and "Enhancing Genetic Medicine: Rapid and Cost-Effective Molecular Diagnosis for a GJB2 Founder Mutation for Hearing Impairment in Ghana" (33) were included based on their focus on digital health and personalised medicine. Studies were included if they met predefined criteria related to the research question, such as specific study designs, populations, interventions, and outcomes. For instance, we included empirical studies that provided evidence-based insights into healthcare practices in low-resource settings, while excluding opinion pieces or studies that did not focus on our primary areas of interest.

In addition to the manual screening process, we integrated an automated selection process to enhance the precision and efficiency of our comprehensive scoping review. We leveraged advanced machine learning techniques, employing Python scripts for preprocessing and classification of relevant studies. Initially, we combined titles and abstracts into a single text field and categorized them using keywords through Naive Bayes, SVM, and Random Forest classifiers, each assessing the data’s context and thematic relevance. To further enhance reliability and reduce potential biases, we implemented an ensemble learning strategy using a “VotingClassifier” that combined the strengths of the individual models. This ensemble approach utilized a ‘hard’ voting mechanism where the majority vote determined the final classification, significantly improving the accuracy of our literature categorization and ensuring a robust review process focused on the most pertinent studies. Each phase was meticulously tested and validated, culminating in a dataset prepared for in-depth analysis (31).

#### Stage 2: Full-Text Review

The full texts of studies that passed the initial screening were retrieved and reviewed in detail. Two reviewers independently assessed these full texts to ensure they met the inclusion criteria. Studies like "Factors Influencing Adoption of eHealth Technologies in Ghana" (34) and "Genomic Medicine Without Borders: Which Strategies Should Be Embraced to Improve Access and Equity?" (35) were included after a detailed review confirmed their relevance. Any discrepancies during the full-text review were resolved through discussion and consensus between the reviewers.

#### Metadata Management

Metadata from the databases were retrieved and uploaded into the Zotero reference management system. This system facilitated the organisation and cleaning of the data. Using Zotero’s features, we cleaned up the metadata and retrieved any missing information. This step included eliminating retracted papers and deleting duplicate records. For example, the retracted paper "The relationship between C-reactive protein and levels of various cytokines in patients with COVID-19: A systematic review and correlation analysis" (36) was identified and removed from the dataset to maintain data integrity. After screening the titles and abstracts for inclusion, the final cleaned metadata was exported for further analysis.

#### Data Extraction and Charting

Data were extracted using a standardised form (Supplementary Table 2) designed to capture key variables such as author, title, year, abstract, study design, setting/context, population, interventions, outcomes, and key findings. The extraction form was pilot-tested and refined to ensure clarity, comprehensiveness, and consistency across all reviewers. Extracted data (Supplementary Table 3) were charted to provide a descriptive summary of the included studies.

## Data Analysis

### Descriptive Statistics

Descriptive statistics were used to summarise study characteristics such as the number of studies, publication years, and interventions, aiding in understanding the distribution and trends within the dataset. The data were also summarised by context and theme, providing insights into the contextual and thematic distribution of the studies. This involved grouping the data based on different contexts (Mostly Ghana and Low resource setting) and themes identified during extraction.

### Thematic Analysis

Thematic analysis was conducted to identify patterns and themes related to the implementation and impact of digital health solutions, personalised medicine, and N-of-1 trials. This process involved analysing the frequency and context of words and phrases in the study abstracts. Common themes were derived by filtering out frequent but less informative words (stopwords) and focusing on the significant terms that appeared frequently across the abstracts. For example, recurring themes such as "mHealth," "healthcare," and “intervention” were identified.

### Quality and Relevance Assessment

The quality of materials was assessed based on their methodology, relevance to the research objectives, and data robustness. Studies were evaluated with a preference for randomised controlled trials, cohort studies, and systematic reviews due to their methodological rigour. However, case studies and qualitative research were also considered if they provided valuable insights. The quality evaluation involved a critical assessment of potential biases, particularly regarding study design, participant selection, and outcome reporting. Each study was assigned a quality score, ranging from 1 (critical flaws) to 5 (excellent), based on how well it met these criteria.

Relevance assessment focused on the alignment of each study with the review objectives, its contextual relevance to low-resource settings like Ghana, and its ability to provide actionable insights and innovative strategies. Studies were also evaluated for their coverage of key themes, such as enhancing healthcare access and quality, applying personalised treatment strategies, and assessing the effectiveness of digital health tools. Additionally, the assessment considered the study’s contribution to filling knowledge gaps and its exploration of socio-cultural and economic factors in the adoption of healthcare innovations. Each study was assigned a relevance score from 1 (minimal relevance) to 5 (maximum relevance) to facilitate prioritisation for inclusion in the review, ensuring that the most pertinent and high-quality studies were selected. See Supplementary Text 2 for details.

## Results

### Overview of Included Studies

An overview of the retrieved and included studies in our scoping review on digital health solutions and personalised medicine in Ghana is given in the PRISMA (Preferred Reporting Items for Systematic Reviews and Meta-Analyses) flow diagram (37) in Figure 1 (see also Supplementary Table 4). The flow diagram shows that out of 6,282 records identified, 59 records were screened, 49 were assessed for eligibility, and 40 new studies were ultimately included, reflecting a systematic and rigorous screening process (38). The automated filtering step of eligible articles removed 4,057 articles, see Supplementary Table 5 for more details.

**Figure 1:**
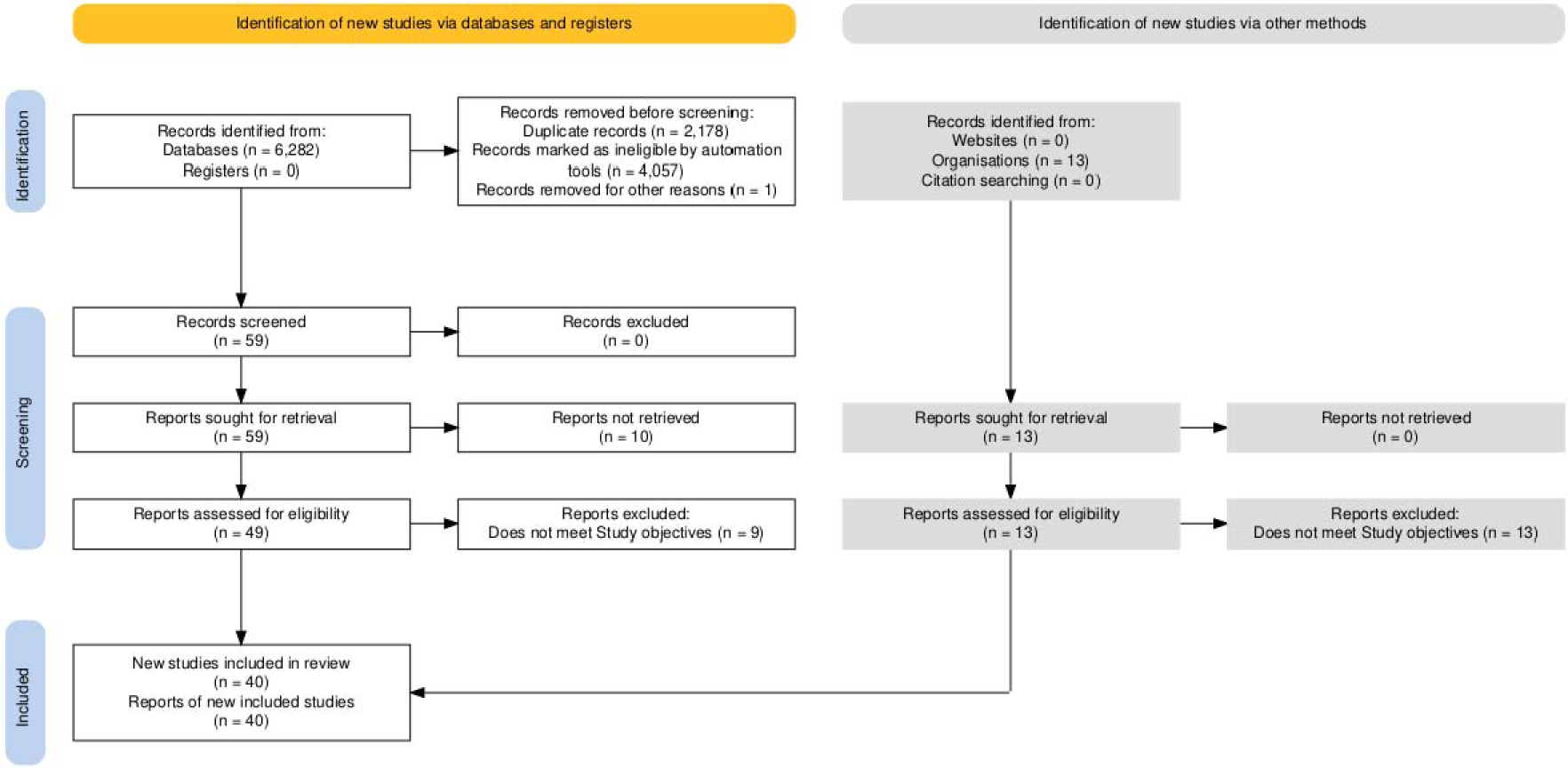
Overview of included Studies.

### Study Characteristics

The distribution of digital health and personalised medicine studies by geographical context and thematic focus reveals significant trends (Figure 2). Most studies (95.6%) are centred on Ghana, indicating a concentrated interest in digital health solutions and personalised medicine within this specific context. In contrast, only 4.4% of the studies focus on broader low-resource settings. The thematic focus is similarly skewed, with 93.3% of the articles concentrating on digital health, while only 6.7% address personalised medicine. These trends suggest that digital health, particularly in Ghana, is an area of interest, reflecting the global push towards leveraging technology to improve healthcare in specific regions (39).

**Figure 2:**
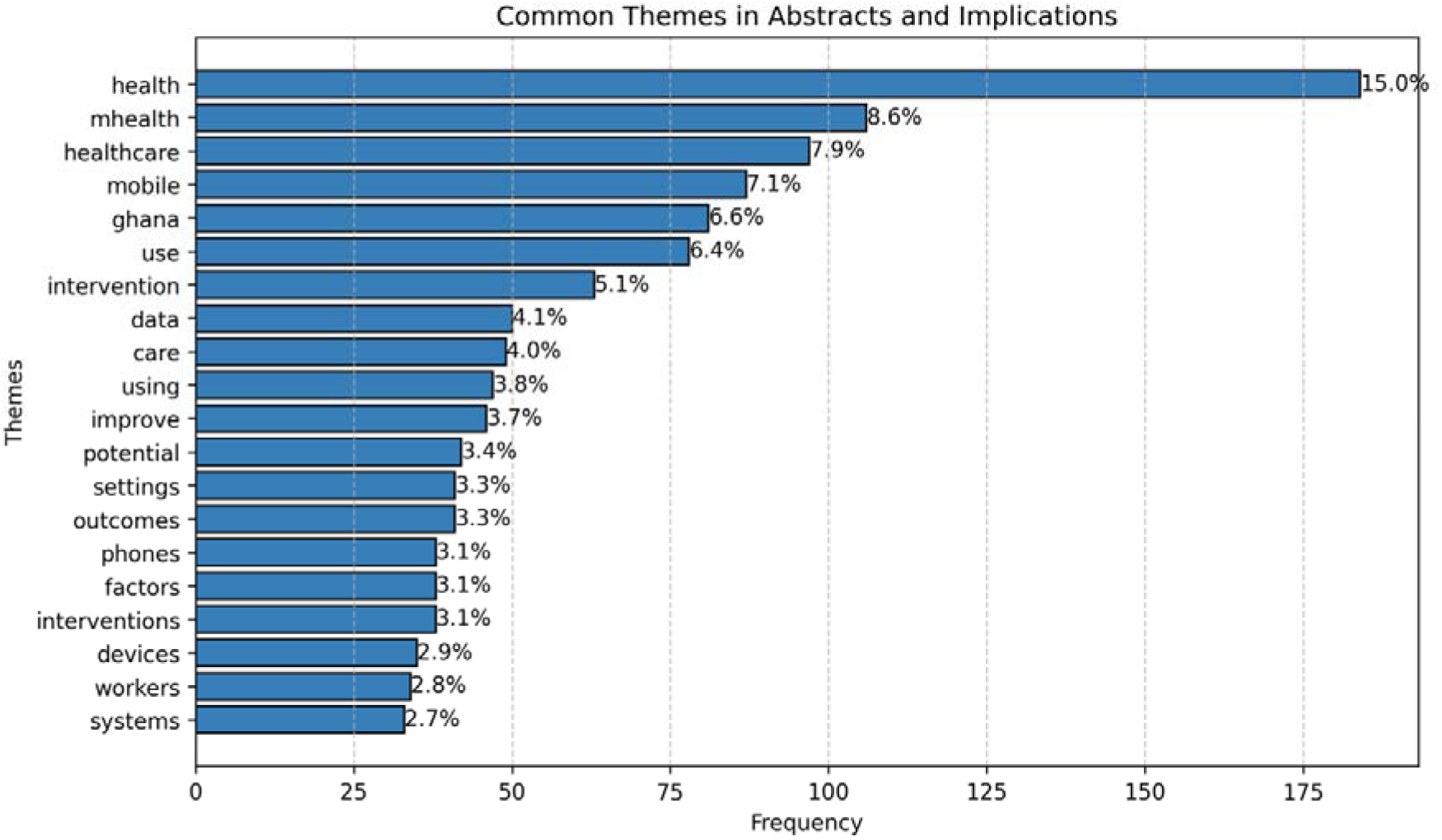
Common themes in titles and abstracts.

Common themes and key terms frequently mentioned in the literature on digital health and personalized medicine highlight the primary areas of focus and interest. Terms like "health," "mhealth," "healthcare," "mobile," and "Ghana" appear most frequently, reflecting the emphasis on mobile health technologies and their implementation in healthcare systems, especially in the Ghanaian context. Other significant terms include "use," "intervention," "data," "care," "improve," and "settings," indicating a broad range of topics related to the practical applications and benefits of digital health solutions. These themes underscore the centrality of mobile health in advancing healthcare delivery and the diverse applications of digital technologies in this field (40). See Supplementary Figures 2-3 for a visual illustration using word clouds.

Publication trends for digital health and personalised medicine research have evolved significantly over time, reflecting changing interests and advancements in the field (Figure 3). Based on the selected papers defined by our parameters (Supplementary Text 1 and 2), the number of publications in Ghana has fluctuated, peaking at 8 publications in 2019, with significant research activity followed by subsequent drops to around 2-3 publications per year. This trend mirrors global interest in digital health technologies, with peaks in publication activity corresponding to advancements and increased funding in these areas. Similarly, digital health research shows significant fluctuations, with a peak of 8 publications in 2019, indicating aligned research interests. Personalized medicine, while showing consistent publication rates with peaks of 2 publications in some years, reflects a growing but still limited focus, highlighting the need for more research in this emerging field (41,42). No studies were included between 2000 and 2016, indicating a late start in research activities in these areas.

**Figure 3:**
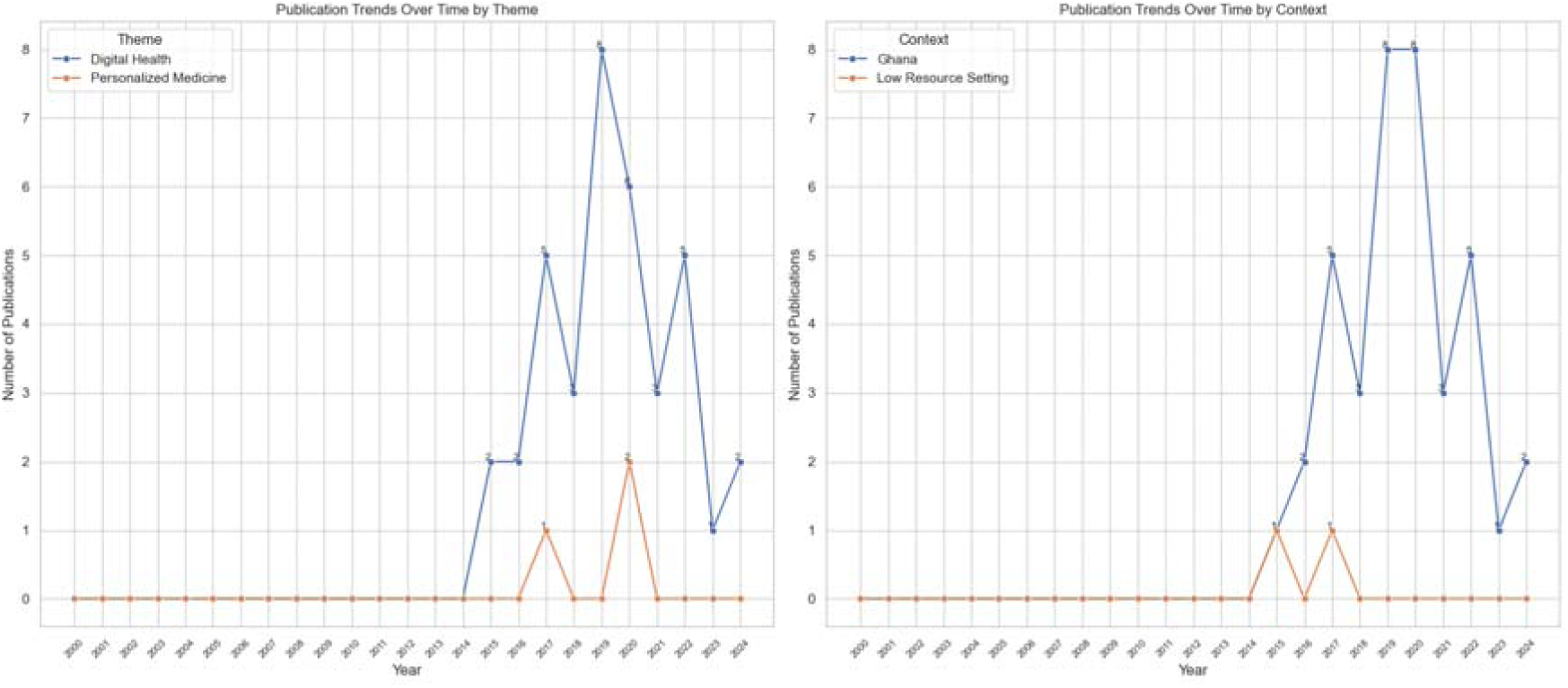
Publication Trends by Theme and Context.

### Types of Interventions

Interventions in digital health and personalised medicine are diverse and categorised into several types, reflecting the broad scope and application of these technologies. The categorisation process involved several steps. First, the "Intervention Type" column from the extracted content served as the primary data source. Each intervention type was reviewed to identify common themes. The identified themes included mHealth solutions, eHealth solutions, genomic medicine, wearable technology, and advanced telemedicine (Figure 4).

**Figure 4:**
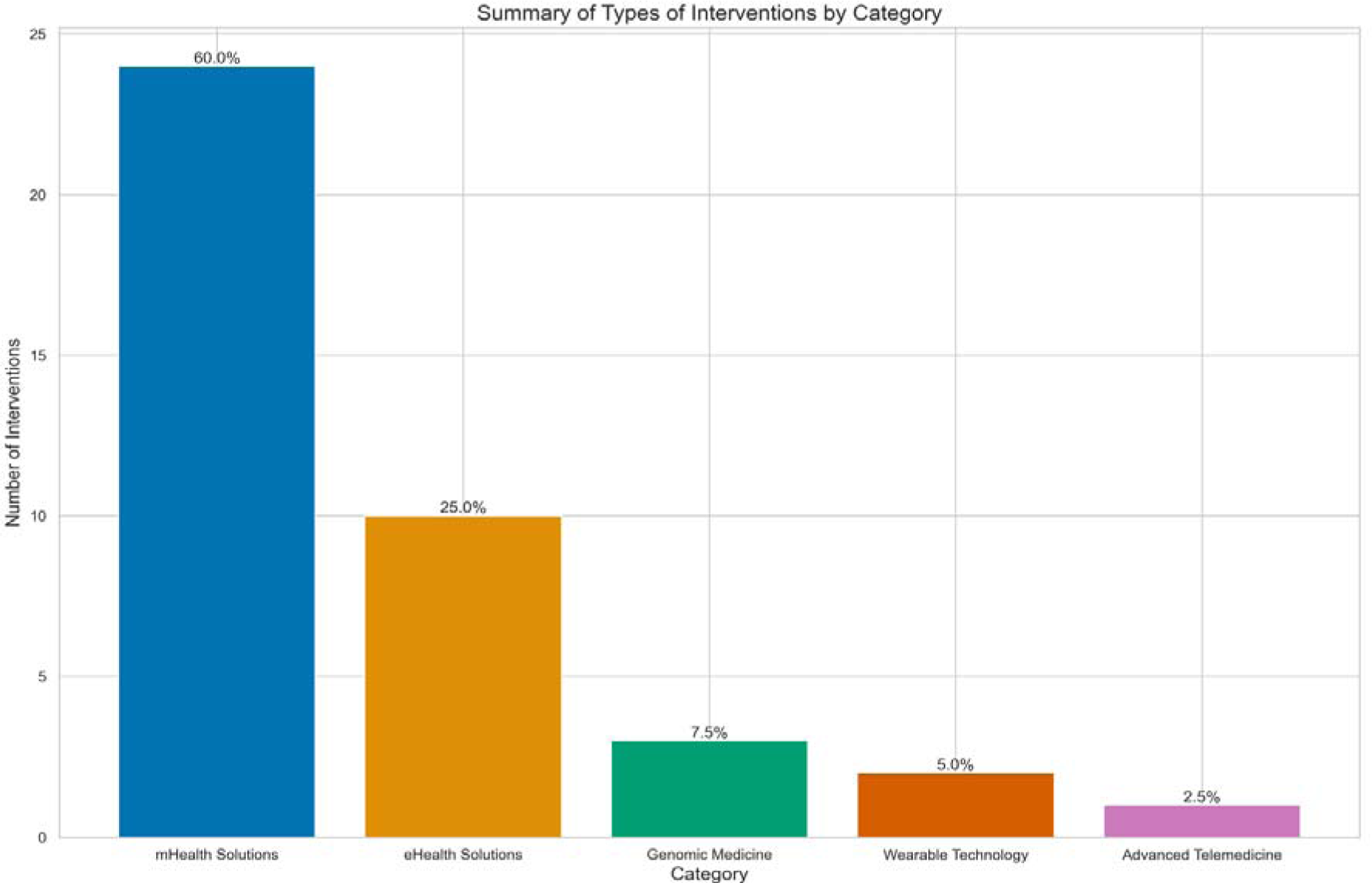
Types of Interventions.

Most interventions (60.0%) are mHealth solutions, indicating a strong focus on mobile health technologies such as clinical decision-making support systems and smartphone-based training programs (33,43,44). For example, the study by Amoakoh et al. (39) explores using mHealth to support clinical decision-making in Ghana, and Osei et al. (45) discuss the availability and use of mobile health technologies for disease surveillance. Additionally, research by Kesse-Tachi et al. (34) investigates factors influencing the adoption of eHealth technologies, including mobile health applications.

eHealth solutions represent 25.0% of interventions, including electronic health records and institutionalising health technology assessments (46,47). The research by Koduah et al. (48) focuses on institutionalising health technology assessments in Ghana, while Abdulai et al. (49) study health providers’ readiness for electronic health records. Another notable study by Opoku et al.(50) examines the sustainability and scale-up of mobile health initiatives, highlighting the importance of integrating eHealth solutions with existing health systems. Asgary et al. (51) delve into the acceptability and implementation challenges of telemedicine in diverse healthcare settings. Furthermore, Kesse-Tachi et al. (34) investigate the factors influencing the adoption of eHealth technologies, providing insights into the barriers and facilitators for effective implementation.

Genomic medicine accounts for 7.5%, focusing on applications like rapid molecular diagnostic tests. For example, Adadey et al. (33) discuss enhancing genetic medicine through rapid and cost-effective diagnostic methods. Wearable technology constitutes 5.0%, reflecting the integration of wearable devices in health interventions. For instance, the study by Agarwal et al. (48) tracks health commodity inventory and notifications using wearable technology. Advanced telemedicine makes up 2.5%, indicating niche but innovative approaches in telemedicine. Examples include research on telemedicine applications and their impacts on healthcare delivery in resource-limited settings (52,53). Agarwal et al. (41) explore decision support tools via mobile devices to improve healthcare delivery, highlighting innovative approaches within advanced telemedicine.

### Quality and Relevance Scores

The quality and relevance of studies in systematic reviews of digital health and personalized medicine literature is assessed using specific scoring systems (Figure 5). Most studies in this review received high scores: 57.8% scored a 5 (the highest quality), and 37.8% scored a 4. Only a small fraction (4.4%) received a quality score of 3, indicating robust methodologies in most studies. Relevance scores are similarly high, with 91.1% of the articles scoring a 5, highlighting their direct applicability and importance to the research objectives. These assessments ensure that the included studies are not only methodologically sound but also highly pertinent to the research questions being addressed (54). Notably, the search for N-of-1 trials returned no results, indicating a gap in the literature regarding this specific type of study.

**Figure 5:**
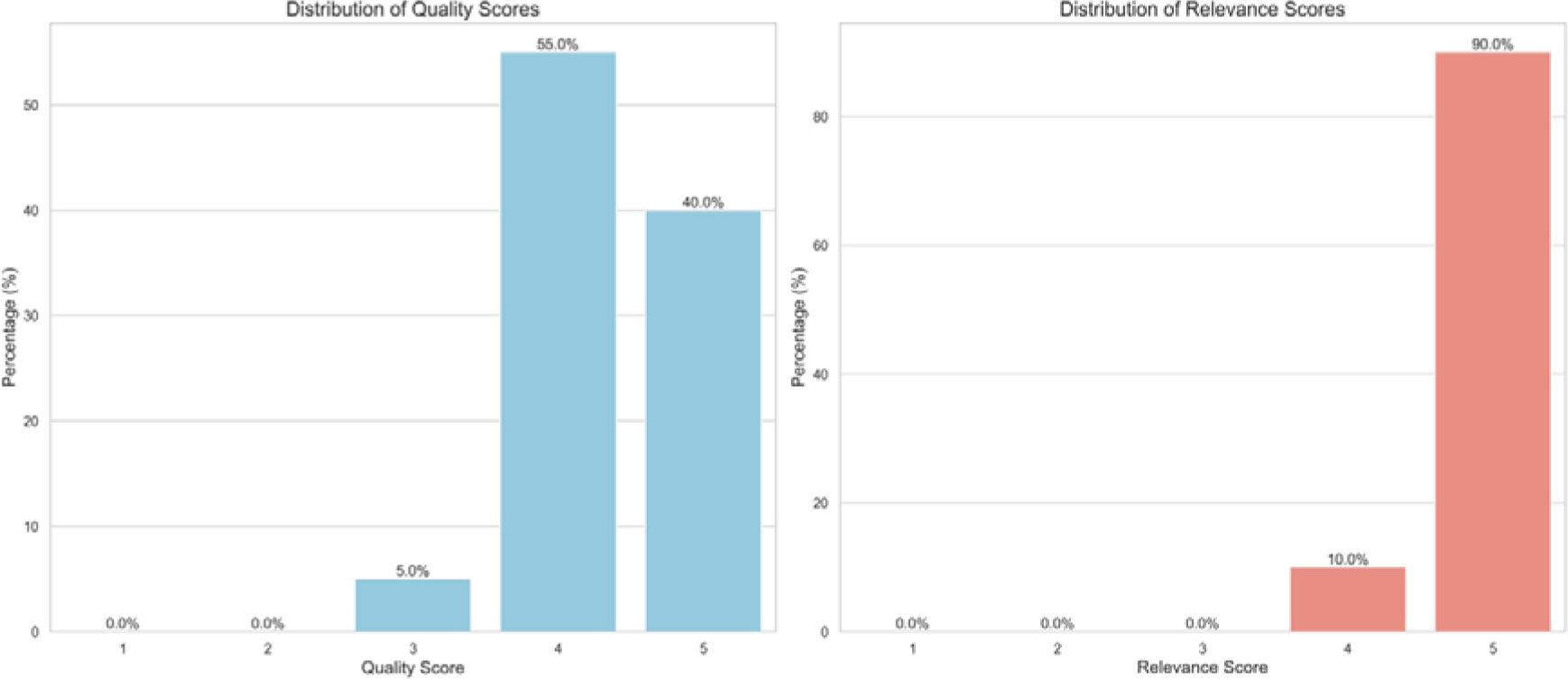
Quality and Relevance Score of Articles.

## Discussion

This scoping review highlights several key findings regarding the implementation and impact of digital health solutions, personalized medicine, and N-of-1 trials in Ghana.

### Publication Trends and Gaps

Publication trends for digital health and personalized medicine research have evolved significantly over time, reflecting changing interests and advancements in the field. Based on the selected papers defined by our parameters (Supplementary Text 1 and 2), the number of publications in Ghana has fluctuated, peaking at 8 publications in 2019, with significant research activity followed by subsequent drops to around 2-3 publications per year. This trend mirrors global interest in digital health technologies, with peaks in publication activity corresponding to advancements and increased funding in these areas. Similarly, digital health research shows significant fluctuations, with a peak of 8 publications in 2019, indicating aligned research interests. Personalized medicine, while showing consistent publication rates with peaks of 2 publications in some years, reflects a growing but still limited focus, highlighting the need for more research in this emerging field (41,42).

A critical observation is the lack of studies between 2000 and 2016, indicating a late start in research activities in these areas. This gap could be attributed to several factors, including limited technological infrastructure, lack of funding, and insufficient awareness of digital health and personalized medicine concepts during that period. The surge in publications post-2016 coincides with global advancements in digital health technologies and increased investment in healthcare innovation. This delay aligns with the global trend, as noted by Karamagi et al. (2022), who observed similar patterns in sub-Saharan Africa due to initial infrastructural and financial constraints (1).

The COVID-19 pandemic significantly impacted research activities, both positively and negatively. While some studies were delayed or cancelled due to restrictions, the pandemic also accelerated the adoption of digital health solutions, as evidenced by the surge in publications focusing on telemedicine and remote healthcare solutions. This trend is supported by Labrique et al. (2018), who highlighted the rapid scaling of digital health technologies during public health crises (17).

The observed delay in the start of research activities in digital health and personalized medicine in Ghana is not unique. Similar trends have been noted globally, particularly in low- and middle-income countries. For example, a study by Fatehi et al. (2020) highlighted that the lack of early research in digital health could be attributed to limited technological infrastructure and funding in these regions (8). The authors noted that many low-resource settings faced significant barriers in adopting and integrating new technologies due to inadequate infrastructure, which is consistent with our findings in Ghana.

The increase in publications post-2016 correlates with global advancements in digital health technologies and increased funding for healthcare innovation. This pattern is evident in a global context, as seen in the study by Mathews et al. (2019), which reported a surge in digital health publications and innovations around this time due to enhanced funding and technological advancements (3). The alignment of publication trends in Ghana with these global patterns suggests that local researchers are increasingly engaging with and contributing to broader technological advancements.

Moreover, the impact of the COVID-19 pandemic on accelerating digital health research is a phenomenon observed worldwide. According to a review by Abul-Husn and Kenny (2019), the pandemic acted as a catalyst for the rapid adoption of digital health technologies, as traditional healthcare systems struggled to cope with the crisis (19). The surge in publications focusing on telemedicine and remote healthcare solutions during the pandemic, as noted in our review, aligns with this global trend, indicating that Ghanaian researchers and healthcare providers adapted quickly to the changing landscape.

### Major Focus on Mobile Health Solutions

The review revealed that mobile health (mHealth) interventions are a major focus in the reviewed studies, particularly in Ghana, where 60% of the interventions are mHealth solutions. These include mobile health technologies such as clinical decision-making support systems and smartphone-based training programs aimed at improving healthcare delivery through mobile platforms. These tools provide remote consultations, patient monitoring, and health education. For instance, mobile applications have significantly supported clinical decision-making and improved maternal and neonatal health outcomes in Ghana. Health workers benefit significantly from these tools, which assist in disease diagnosis and treatment support (55). Despite some challenges, the acceptability and implementation of mHealth interventions demonstrate their potential to enhance healthcare accessibility and efficiency (56,57). However, sustainability and scale-up of mHealth interventions require continuous funding and technological support (58).

### Significant Role of Electronic Health Records (EHRs)

Electronic Health Records (EHRs) represent another significant intervention, accounting for 24% of the reviewed studies. EHRs have been shown to improve data management, enhance patient care coordination, and facilitate better health outcomes. Abdulai and Lee (46,49), highlighted the readiness of health providers for EHR implementation, noting the considerable benefits in data management and patient care oversight. However, they also identified challenges such as the cost of implementation and resistance to change among healthcare providers, which need to be addressed for successful adoption. Factors influencing the adoption of eHealth technologies, including EHRs, underline the necessity of user training and ongoing support (34).

### Limited Integration of Personalized Medicine

The integration of personalized medicine in Ghana remains limited, with only a small fraction of the studies focusing on this area. Personalized medicine has the potential to tailor healthcare based on individual genetic, environmental, and lifestyle factors, thereby improving patient outcomes. However, the current infrastructure and resources in Ghana pose significant challenges to the widespread adoption of personalized medicine. Addressing these challenges requires substantial investment in healthcare infrastructure, education for healthcare providers, and the development of policies to support personalized approaches to medicine (33,35).

### Lack of N-of-1 Trials

Despite the transformative potential of N-of-1 trials for individualized patient care in Ghana, this review found no studies specifically focused on these trials or similar personalized trials within the Ghanaian context. This absence underscores a critical need for initiatives to build both awareness and capacity for implementing N-of-1 methodologies within the healthcare system. Enhancing healthcare providers’ understanding of this design, along with establishing technical support for trial implementation, are essential steps toward integrating N-of-1 trials into routine clinical practice in Ghana. By addressing these gaps, N-of-1 trials could become a powerful tool for advancing personalized healthcare in low-resource settings like Ghana. Then, digital open-source tools could be used that have been made available for designing and conducting N-of-1 trials in order to integrate citizens and patients into evaluating the effectiveness of health interventions for them directly to personalize prevention or treatment (59).

### Limitations of the Study

The scoping review presents several limitations that should be acknowledged to provide context for the findings and interpretations. First, many of the included studies focus on Ghana, with only a small fraction addressing broader low-resource settings. This may be partly caused by our search terms which did not specifically include other countries in low-resource settings by name, which might limit the generalizability of the findings to other similar contexts within Sub-Saharan Africa or other low-resource regions (45,60,61). Second, the review only included studies published in English, potentially leading to the exclusion of relevant studies published in other languages and introducing a language bias. Additionally, we focused more on peer-reviewed articles which may introduce publication bias, as studies with statistically significant findings are more likely to be published. Third, the included studies vary in their methodological rigor, quality of data, and reporting standards, affecting the reliability and comparability of the findings. For instance, some studies might have limited sample sizes or inadequate controls, impacting the strength of the evidence (51,62). Fourth, the review highlights a notable gap in publications before 2016, which could be due to delayed reporting and publication processes, meaning that earlier digital health interventions and their outcomes might not be fully captured in the review. Finally, the findings indicate challenges related to technological infrastructure and resource availability in Ghana, which can affect the implementation and scalability of digital health interventions, thereby limiting their potential impact (50,63).

### Comparison with Other Studies

The trends observed in Ghana align with global patterns in digital health adoption. For instance, Amoakoh et al. (55) noted a significant increase in the use of mobile health interventions to support clinical decision-making in Ghana, reflecting similar trends in other low-resource settings. Globally, the adoption of mobile health (mHealth) technologies has seen exponential growth (64). Similarly, the use of tracking health commodity inventory and notifications via mobile devices, as discussed by Agarwal et al. (48), highlights the global trend of leveraging mobile health technologies for improving healthcare delivery. This trend is further supported by research from Gandapur et al. (65), which emphasizes the role of mHealth in enhancing health system efficiencies and patient outcomes globally.

The predominance of mHealth solutions in the reviewed studies is consistent with findings from other low-resource settings. For instance, Labrique et al. (17) highlighted the effectiveness of mHealth interventions in improving healthcare delivery in low-and middle-income countries. A report by GSMA (66) indicates that the proliferation of mobile technology has enabled innovative health solutions that cater to the specific needs of underserved populations globally. This global trend underscores the importance of mobile technology as a critical tool for health interventions in resource-limited settings. Additional studies, such as those by Feroz et al. (67) and Holeman et al. (68), have also shown the transformative impact of mHealth solutions in improving maternal and child health in South Asia and Sub-Saharan Africa, respectively.

The review’s findings on the significant role of Electronic Health Records (EHRs) in Ghana are comparable to studies in other regions. For example, Abdulai et al. (49) discussed the readiness of health providers for EHR implementation, noting the benefits and challenges like those observed in other geographical contexts. This is echoed by research from Kruse et al. (69), which found that EHRs improve care coordination, patient safety, and clinical outcomes across diverse healthcare settings. Additionally, a study by Irizarry and Barton (70) highlights the broader adoption of EHRs, emphasizing the need for a sociotechnical approach to fully realize the benefits of EHR implementation. These findings indicate that while challenges exist, the adoption of EHRs is a critical step towards modernizing health systems globally.

Furthermore, Kesse-Tachi et al. (34) and Agarwal et al. (41) explored the factors influencing the adoption of eHealth technologies and decision support tools via mobile devices, respectively, which resonate with the technological and resource constraints identified globally. For example, a study by Gellert et al. (71) found that financial incentives and strong leadership support were crucial for the successful implementation of health IT systems in hospitals across the United States. Additionally, the studies by Adadey et al. (33) and Asgary et al. (51) further corroborate these findings by highlighting the challenges and successes in implementing digital health solutions and personalized medicine in low-resource settings. Research by Greenhalgh et al. (72) also supports the notion that understanding local contexts and involving end-users in the design and implementation process are key to the successful adoption of eHealth technologies.

## Conclusion

This scoping review underscores the significant interest and potential of digital health solutions in Ghana, particularly through mHealth technologies and EHRs. While these technologies have shown considerable promise, their successful implementation requires addressing challenges related to infrastructure and resistance to change. Personalized medicine and N-of-1 trials offer substantial potential for individualized patient care, but their integration into the healthcare system remains limited and requires further exploration. Enhancing digital infrastructure, providing comprehensive training for healthcare providers, and developing supportive policies are crucial steps towards fully realizing the benefits of these innovations in Ghana’s healthcare system. We recommend further research into sustainable models for integrating digital health and N-of-1 trials, especially focusing on creating cost-effective and adaptable trial methodologies suitable for Ghana’s healthcare infrastructure.

## Data Availability

All data produced in the present work are contained in the manuscript.

## Conflicts of Interest

The authors declare no conflicts of interest.

## Abbreviations

EHRs: Electronic Health Records
mHealth: Mobile Health
IT: Information Technology
ICT: Information and Communications Technology
UHC: Universal Health Coverage
SMS: Short Message Service
HIT: Health Information Technology
PRISMA: Preferred Reporting Items for Systematic Reviews and Meta-Analyses
SVM: Support Vector Machine
DOI: Digital Object Identifier
RCTs: Randomized Controlled Trials
AJOL: African Journals Online
CPOE: Computerized Physician Order Entry
NIH: National Institutes of Health
WHO: World Health Organization
CDC: Centers for Disease Control and Prevention
NGOs: Non-Governmental Organizations
GSMA: GSM Association

## Author Contributions

GS and SK generated the review idea and developed the review protocol. GS ran the searches, screened the records, extracted the data, mapped the data and developed the manuscript draft, under supervision of SK. All co-authors critically reviewed the manuscript and approved the final version before submission.

## Notes

### Competing Interest Statement

The authors have declared no competing interest.

### Funding Statement

This study did not receive any funding

