## Supplementary Material for "Exploring Digital Health Solutions: Personalised Medicine and N-of-1 Trials in Ghana: A Scoping Review"

### Supplementary Text 1: Details on Literature Search Queries

**Overview**

This document outlines a comprehensive keyword and phrase strategy for conducting a literature search on Personalised medicine, digital health solutions, N-of-1 trials, and their implementation and challenges in low-resource settings, with a specific focus on Ghana.

**1. Keywords relevant for Personalised Medicine and Precision Medicine**

- Bio individuality
- Personalised Health Care
- Personalised Therapeutics
- Personalised Treatment Plans
- Personal Genomics
- Precision Medicine
- Individualized Therapy
- Tailored Treatment
- Genetic Medicine
- Genomic Medicine
- Pharmacogenomics
- Customized Healthcare
- Stratified Medicine
- Customized Medical Care
- Biomarker-based Treatment

**2. Keywords relevant for Digital Health**

- Health ICT Information and Communication Technology
- Digital Therapeutics
- Health Application Platforms
- Smart Health
- Digital Patient Management
- eHealth
- mHealth Mobile Health
- Telehealth
- Telemedicine
- Health Informatics
- Digital Health Solutions
- Health Technology
- Wearable Devices
- Remote Monitoring
- Electronic Health Records EHR
- Health Apps

- Digital Medicine
- Health Information Technology
- E-Prescribing
- Virtual Health Care

**3. Keywords relevant for N-of-1 Trials**

- Personalised Research Trials
- Individualized Health Trials
- Patient-Centered Trials
- Therapeutic Personalization
- N-of-1 Trials
- Single-Subject Research
- Individualized Trials
- Case-Specific Trials
- Personalised Clinical Trials
- Single-Case Experiments
- Single-Patient Trials
- Individualized Evidence
- Personalised Clinical Decision Making

**Database Sources**

i. PubMed
ii. IEEE Xplore
iii. The Cochrane Library
iv. Google Scholar
v. Government Websites – Mnistry of Health (MoH), Ghana Health Service (GHS)
vi. Directory of Open Access Journals
vii. Other Organizations WHO, CDC, NGOs

**Search Queries**

**Personalised Medicine and Precision Medicine in Low Resource Settings**

"Bio Individuality" AND ("Low-Resource Settings" OR "Developing Countries")
"Personalised Health Care" AND ("Low-Resource Settings" OR "Developing Countries")
"Personalised Therapeutics" AND ("Low-Resource Settings" OR "Developing Countries")
"Personalised Treatment Plans" AND ("Low-Resource Settings" OR "Developing Countries")
"Personal Genomics" AND ("Low-Resource Settings" OR "Developing Countries")
"Precision Medicine" AND ("Low-Resource Settings" OR "Developing Countries")
"Individualized Therapy" AND ("Low-Resource Settings" OR "Developing Countries")
"Tailored Treatment" AND ("Low-Resource Settings" OR "Developing Countries")
"Genetic Medicine" AND ("Low-Resource Settings" OR "Developing Countries")
"Genomic Medicine" AND ("Low-Resource Settings" OR "Developing Countries")
"Pharmacogenomics" AND ("Low-Resource Settings" OR "Developing Countries")
"Customized Healthcare" AND ("Low-Resource Settings" OR "Developing Countries")
"Stratified Medicine" AND ("Low-Resource Settings" OR "Developing Countries")
"Customized Medical Care" AND ("Low-Resource Settings" OR "Developing Countries")

"Biomarker-based Treatment" AND ("Low-Resource Settings" OR "Developing Countries")

("Bio Individuality" OR "Personalised Health Care" OR "Personalised Therapeutics" OR "Personalised Treatment Plans" OR "Personal Genomics" OR "Precision Medicine" OR "Individualized Therapy" OR "Tailored Treatment" OR "Genetic Medicine" OR "Genomic Medicine" OR "Pharmacogenomics" OR "Customized Healthcare" OR "Stratified Medicine" OR "Customized Medical Care" OR "Biomarker-based Treatment") AND ("Low-Resource Settings" OR "Developing Countries")

**Personalised Medicine and Precision Medicine in Ghana**

"Bio Individuality" AND Ghana
"Personalised Health Care" AND Ghana
"Personalised Therapeutics" AND Ghana
"Personalised Treatment Plans" AND Ghana
"Personal Genomics" AND Ghana
"Precision Medicine" AND Ghana
"Individualized Therapy" AND Ghana
"Tailored Treatment" AND Ghana
"Genetic Medicine" AND Ghana
"Genomic Medicine" AND Ghana
"Pharmacogenomics" AND Ghana
"Customized Healthcare" AND Ghana
"Stratified Medicine" AND Ghana
"Customized Medical Care" AND Ghana
"Biomarker-based Treatment" AND Ghana

("Bio Individuality" OR "Personalised Health Care" OR "Personalised Therapeutics" OR "Personalised Treatment Plans" OR "Personal Genomics" OR "Precision Medicine" OR "Individualized Therapy" OR "Tailored Treatment" OR "Genetic Medicine" OR "Genomic Medicine" OR "Pharmacogenomics" OR "Customized Healthcare" OR "Stratified Medicine" OR "Customized Medical Care" OR "Biomarker-based Treatment") AND Ghana

**Digital Health in Low Resource Settings**

"Health ICT" AND ("Low-Resource Settings" OR "Developing Countries")

"Information and Communication Technology" AND ("Low-Resource Settings" OR "Developing Countries")

"Digital Therapeutics" AND ("Low-Resource Settings" OR "Developing Countries")

"Health Application Platforms" AND ("Low-Resource Settings" OR "Developing Countries")

"Smart Health" AND ("Low-Resource Settings" OR "Developing Countries")

"Digital Patient Management" AND ("Low-Resource Settings" OR "Developing Countries")

"eHealth" AND ("Low-Resource Settings" OR "Developing Countries")

"mHealth" AND ("Low-Resource Settings" OR "Developing Countries")

"Mobile Health" AND ("Low-Resource Settings" OR "Developing Countries")

"Telehealth" AND ("Low-Resource Settings" OR "Developing Countries")

"Telemedicine" AND ("Low-Resource Settings" OR "Developing Countries")

"Health Informatics" AND ("Low-Resource Settings" OR "Developing Countries")

"Digital Health Solutions" AND ("Low-Resource Settings" OR "Developing Countries")

"Health Technology" AND ("Low-Resource Settings" OR "Developing Countries")

"Wearable Devices" AND ("Low-Resource Settings" OR "Developing Countries")

"Remote Monitoring" AND ("Low-Resource Settings" OR "Developing Countries")

"Electronic Health Records" AND ("Low-Resource Settings" OR "Developing Countries")

"EHR" AND ("Low-Resource Settings" OR "Developing Countries")

"Health Apps" AND ("Low-Resource Settings" OR "Developing Countries")

"Digital Medicine" AND ("Low-Resource Settings" OR "Developing Countries")

"Health Information Technology" AND ("Low-Resource Settings" OR "Developing Countries")

"E-Prescribing" AND ("Low-Resource Settings" OR "Developing Countries")

"Virtual Health Care" AND ("Low-Resource Settings" OR "Developing Countries")

("Health ICT" OR "Information and Communication Technology" OR "Digital Therapeutics" OR "Health Application Platforms" OR "Smart Health" OR "Digital Patient Management" OR "eHealth" OR "mHealth" OR "Mobile Health" OR "Telehealth" OR "Telemedicine" OR "Health Informatics" OR "Digital Health Solutions" OR "Health Technology" OR "Wearable Devices" OR "Remote Monitoring" OR "Electronic Health Records" OR "EHR" OR "Health Apps" OR "Digital Medicine" OR "Health Information Technology" OR "E-Prescribing" OR "Virtual Health Care") AND ("Low-Resource Settings" OR "Developing Countries")

**Digital Health in Ghana**

"Health ICT" AND Ghana
"Information and Communication Technology" AND Ghana
"Digital Therapeutics" AND Ghana
"Health Application Platforms" AND Ghana
"Smart Health" AND Ghana
"Digital Patient Management" AND Ghana
"eHealth" AND Ghana
"mHealth" AND Ghana
"Mobile Health" AND Ghana
"Telehealth" AND Ghana
"Telemedicine" AND Ghana
"Health Informatics" AND Ghana
"Digital Health Solutions" AND Ghana
"Health Technology" AND Ghana
"Wearable Devices" AND Ghana
"Remote Monitoring" AND Ghana
"Electronic Health Records" AND Ghana
"EHR" AND Ghana
"Health Apps" AND Ghana
"Digital Medicine" AND Ghana
"Health Information Technology" AND Ghana
"E-Prescribing" AND Ghana
"Virtual Health Care" AND Ghana

("Health ICT" OR "Information and Communication Technology" OR "Digital Therapeutics" OR "Health Application Platforms" OR "Smart Health" OR "Digital Patient Management" OR "eHealth" OR "mHealth" OR "Mobile Health" OR "Telehealth" OR "Telemedicine" OR "Health Informatics" OR "Digital Health Solutions" OR "Health Technology" OR "Wearable Devices" OR "Remote Monitoring" OR "Electronic Health Records" OR "EHR" OR "Health Apps" OR "Digital Medicine" OR "Health Information Technology" OR "E-Prescribing" OR "Virtual Health Care") AND Ghana

### Supplementary Text 2: Quality and Relevance Rating of Literature Search

**Quality Assessment Notes**

a. Quality evaluation based on methodology, relevance to the research objectives, and data robustness.
b. Preference for randomized controlled trials, cohort studies, and systematic reviews. Case studies and qualitative research are included if they provide significant insights.
c. Relevance to Ghana/Low-Resource Settings: Necessity for studies to offer pertinent insights into healthcare in resource-limited, socio-economically challenging settings like Ghana.
d. Implications for Personalized Medicine and Digital Health: Literature must contribute to the advancement of personalized medicine and digital health, with applicability in real-world settings.
e. Risk of Bias Evaluation: Critical assessment of potential biases, particularly regarding study design, participant selection, and outcome reporting.

**Quality Score/Rating**

1 (Fail): Critical flaws invalidate the results.
2 (Poor): Significant shortcomings in quality and methodology.
3 (Fair): Meets some quality criteria; limitations may affect result reliability.
4 (Good): Meets most quality criteria; some limitations do not compromise results.
5 (Excellent): Meets all quality criteria with minor limitations.

**Relevance Assessment Notes**

1. Alignment with Review Objectives: Studies must directly contribute to the understanding of personalized medicine and digital health solutions, particularly in the context of Ghana or similar low-resource settings. The degree to which a study's focus, findings, and implications support the review's specific aims will be critically assessed.
2. Contextual Relevance: The extent to which the research addresses the unique challenges, opportunities, and needs of low-resource settings is essential. Studies that provide insights into healthcare practices, barriers to technology adoption, patient engagement strategies, and the implementation of N-of-1 trials in such environments will be prioritized.
3. Insightfulness and Actionability: Preference will be given to literature that offers actionable insights, innovative strategies, or evidence-based recommendations that can inform the development, deployment, and scaling of personalized medicine and digital health interventions in Ghana and comparable contexts.
4. Coverage of Key Themes: Research must cover one or more of the key themes of the review, including but not limited to the role of technology in enhancing healthcare access and quality, the application of personalized treatment strategies, and the evaluation of digital health tools' effectiveness.
5. Contribution to Knowledge Gaps: Studies that address identified gaps in the literature or introduce new perspectives on the integration of personalized medicine and digital health solutions in low-resource settings will be highly valued. The ability of a study to fill these gaps will be a critical factor in its relevance assessment.

6. Socio-Cultural and Economic Considerations: Given the significance of socio-cultural and economic factors in the adoption and impact of healthcare innovations, studies that explore these aspects within the context of personalized medicine and digital health are considered highly relevant.

**Relevance Scoring Process**

Each study will be assigned a Relevance Score from 1 to 5, based on how well it meets the above criteria, with 1 indicating minimal relevance and 5 indicating maximum relevance to the project's objectives. This scoring will facilitate the prioritization of literature for inclusion in the review.

**Relevance Score (1-5)**

1: Study focus significantly diverges from project objectives.
2: Limited application of study focusses to project objectives.
3: Moderate relevance to project objectives.
4: Relevant study providing useful data towards project objectives.
5: Directly addresses project objectives with high-value insights.

### Supplementary Tables

| SECTION | ITEM | PRISMA-ScR CHECKLIST ITEM | REPORTED ON PAGE # |
|---|---|---|---|
| **TITLE** | | | |
| Title | 1 | Identify the report as a scoping review. | 1 |
| **ABSTRACT** | | | |
| Structured summary | 2 | Provide a structured summary that includes (as applicable): background, objectives, eligibility criteria, sources of evidence, charting methods, results, and conclusions that relate to the review questions and objectives. | 1 |
| **INTRODUCTION** | | | |
| Rationale | 3 | Describe the rationale for the review in the context of what is already known. Explain why the review questions/objectives lend themselves to a scoping review approach. | 1 |
| Objectives | 4 | Provide an explicit statement of the questions and objectives being addressed with reference to their key elements (e.g., population or participants, concepts, and context) or other relevant key elements used to conceptualize the review questions and/or objectives. | 1 |
| **METHODS** | | | |
| Protocol and registration | 5 | Indicate whether a review protocol exists; state if and where it can be accessed (e.g., a Web address); and if available, provide registration information, including the registration number. | 6 |
| Eligibility criteria | 6 | Specify characteristics of the sources of evidence used as eligibility criteria (e.g., years considered, language, and publication status), and provide a rationale. | 6 |
| Information sources* | 7 | Describe all information sources in the search (e.g., databases with dates of coverage and contact with authors to identify additional sources), as well as the date the most recent search was executed. | 6 |
| Search | 8 | **Present the full electronic search strategy for at least 1 database, including any limits used, such that it could be repeated.** | 6-7 |
| Selection of sources of evidence† | 9 | State the process for selecting sources of evidence (i.e., screening and eligibility) included in the scoping review. | 6-7 |

| SECTION | ITEM | PRISMA-ScR CHECKLIST ITEM | REPORTED ON PAGE # |
|---|---|---|---|
| Data charting process‡ | 10 | Describe the methods of charting data from the included sources of evidence (e.g., calibrated forms or forms that have been tested by the team before their use, and whether data charting was done independently or in duplicate) and any processes for obtaining and confirming data from investigators. | 7 |
| Data items | 11 | List and define all variables for which data were sought and any assumptions and simplifications made. | 6-7 |
| Critical appraisal of individual sources of evidence§ | 12 | If done, provide a rationale for conducting a critical appraisal of included sources of evidence; describe the methods used and how this information was used in any data synthesis (if appropriate). | 7 |
| Synthesis of results | 13 | Describe the methods of handling and summarizing the data that were charted. | 7 |
| **RESULTS** | | | |
| Selection of sources of evidence | 14 | Give numbers of sources of evidence screened, assessed for eligibility, and included in the review, with reasons for exclusions at each stage, ideally using a flow diagram. | 9 |
| Characteristics of sources of evidence | 15 | For each source of evidence, present characteristics for which data were charted and provide the citations. | 9-10 |
| Critical appraisal within sources of evidence | 16 | If done, present data on critical appraisal of included sources of evidence (see item 12). | 33-44 |
| Results of individual sources of evidence | 17 | For each included source of evidence, present the relevant data that were charted that relate to the review questions and objectives. | 45-46 |
| Synthesis of results | 18 | Summarize and/or present the charting results as they relate to the review questions and objectives. | 9-12, 38 |
| **DISCUSSION** | | | |
| Summary of evidence | 19 | Summarize the main results (including an overview of concepts, themes, and types of evidence available), link to the review questions and objectives, and consider the relevance to key groups. | 13-15 |
| Limitations | 20 | Discuss the limitations of the scoping review process. | 15 |
| Conclusions | 21 | Provide a general interpretation of the results with respect to the review questions and | 13-15 |

| SECTION | ITEM | PRISMA-ScR CHECKLIST ITEM | REPORTED ON PAGE # |
|---|---|---|---|
| | | objectives, as well as potential implications and/or next steps. | |
| **FUNDING** | | | |
| Funding | 22 | Describe sources of funding for the included sources of evidence, as well as sources of funding for the scoping review. Describe the role of the funders of the scoping review. | No funding to report. |

**Supplementary Table 1:** Preferred Reporting Items for Systematic reviews and Meta-Analyses extension for Scoping Reviews (PRISMA-ScR) Checklist

| Keyword | Author | Title | Abstract | Year | DOI | URL | Theme | Source | Full Text Available (Y/N) | Study Design | Study Setting/Context | Study Population | Intervention/Exposure | Comparison | Primary Outcomes | Secondary Outcomes | Key Findings | Quality Assessment Notes | Relevance to Ghana/Low-Resource Settings | Implications for Personalized Medicine and Digital Health | Risk of Bias | Additional Notes | Relevance Score (1-5) | Quality Score/Rating (1-5) | Final Inclusion (Y/N) |
|---|---|---|---|---|---|---|---|---|---|---|---|---|---|---|---|---|---|---|---|---|---|---|---|---|---|
| unique identifier | Main authors | Title of the paper | abstract | Publication year | DOI | access link | major theme of the paper | database or journal where paper was accessed | whether full document is available or not | Description of the study design | Location and setting of the study | Description of the participants | Details of what is being tested or observed | What the main intervention is being compared to | The main outcomes being measured | Any secondary outcomes being measured | Summary of the study's results | Notes on quality and bias | How the study is relevant to low-resource settings | How the findings relate to personalized medicine and digital health | Assessed level of bias | Any other relevant notes | Score from 1 to 5 on relevance | Score or rating on quality | Yes/No if included in the review |
| 1 | | | | | | | | | | | | | | | | | | | | | | | | | |
| 2 | | | | | | | | | | | | | | | | | | | | | | | | | |
| 3 | | | | | | | | | | | | | | | | | | | | | | | | | |

**Supplementary Table 2. Data Extraction Forms**

This table is a comprehensive data extraction template for a literature review, cataloguing key study details such as bibliographic information, outcomes, and relevance to personalized medicine and low-resource settings

| Sn | Author | Title | Year | Theme | Context |
|---|---|---|---|---|---|
| 1 | Amoakoh HB, Ansah EK., Klipstein-Grobusch K, Grobbee DE, Amoakoh-Coleman M, Oduro-Mensah E, Sarpong C, Frimpong E, Kayode GA, Agyepong IA | Using Mobile Health to Support Clinical Decision-Making to Improve Maternal and Neonatal Health Outcomes in Ghana: Insights of Frontline Health Worker Information Needs | 2019 | Digital Health | Ghana |
| 2 | Osei E, Mashamba-Thompson TP., Agyei K, Tlou B | Availability and Use of Mobile Health Technology for Disease Diagnosis and Treatment Support by Health Workers in the Ashanti Region of Ghana: A Cross-Sectional Survey | 2021 | Digital Health | Ghana |
| 3 | Agarwal, S, S; Glenton, C; Henschke, N; Tamrat, T; Bergman, MS; Mehl, GL; Lewin | Tracking health commodity inventory and notifying stock levels via mobile devices: a mixed methods systematic review | 2020 | Digital Health | Ghana |

| | | | | | |
|---|---|---|---|---|---|
| 4 | Koduah, Justice, Augustina; Boadi, Jessica Anim; Azeez, Joycelyn Naa Korkoi; Asare, Brian Adu; Yevutsey, Saviour; Gyansa-Lutterodt, Martha; Nonvignon | Institutionalizing Health Technology Assessment in Ghana: Enablers, Constraints, and Lessons | 2023 | Digital Health | Ghana |
| 5 | Abdulai, Fuseini, Abdul-Fatawu; Adam | Health providers' readiness for electronic health records adoption: A cross-sectional study of two hospitals in northern Ghana. | 2020 | Digital Health | Ghana |
| 6 | Kesse-Tachi, Ebenezer, Agyenna; Asmah, Alexander Ekow; Agbozo | Factors influencing adoption of eHealth technologies in Ghana | 2019 | Digital Health | Ghana |
| 7 | Agarwal, S, S; Glenton, C; Tamrat, T; Henschke, N; Maayan, N; F, MS; Mehl, GL; Lewin | Decisionsupport tools via mobile devices to improve quality of care in primary healthcare settings | 2021 | Digital Health | Ghana |
| 8 | Asgary R, Adanu R., Cole H, Adongo P, Nwameme A, Maya E, Adu-Amankwah A, Barnett H | Acceptability and implementation challenges of smartphone-based training of community health nurses for visual inspection with acetic acid in Ghana: mHealth | 2019 | Digital Health | Ghana |

| | | | | | |
|---|---|---|---|---|---|
| | | and cervical cancer screening | | | |
| 9 | Opoku D, Quentin W., Busse R | Achieving Sustainability and Scale-Up of Mobile Health Noncommunicable Disease Interventions in Sub-Saharan Africa: Views of Policy Makers in Ghana | 2019 | Digital Health | Ghana |
| 10 | Adadey, Ambroise, Samuel M.; Wonkam, Edmond Tingang; Aboagye, Elvis Twumasi; Quansah, Darius; Asante-Poku, Adwoa; Quaye, Osbourne; Amedofu, Geoffrey K.; Awandare, Gordon A.; Wonkam | Enhancing Genetic Medicine: Rapid and Cost-Effective Molecular Diagnosis for an iGJB2i Founder Mutation for Hearing Impairment in Ghana | 2020 | Personalized Medicine | Ghana |

| | | | | | |
|---|---|---|---|---|---|
| 11 | Amoakoh HB, Ansah EK., Klipstein-Grobusch K, Amoakoh-Coleman M, Agyepong IA, Kayode GA, Sarpong C, Grobbee DE | The effect of a clinical decision-making mHealth support system on maternal and neonatal mortality and morbidity in Ghana: study protocol for a cluster randomized controlled trial | 2017 | Digital Health | Ghana |
| 12 | Lo S, Cutler B; 3DTM (3D Telemedicine) Collaborative Research Group., Rose A, Fowers S, Darko K, Britto A, Spina T, Ankrah L, Godonu A, Ntreh D, Lalwani R, Graham C, Tittsworth D, McIntyre A, O'Dowd C, Watson S, Maguire R, Hoak A, Ampomah O | Ghana 3D Telemedicine International MDT: A proof-of-concept study | 2024 | Digital Health | Ghana |
| 13 | Peprah, Paulinus, Prince; Abalo, Emmanuel Mawuli; Agyemang-Duah, Williams; Gyasi, Razak M.; Reforce, Okwei; Nyonyo, Julius; Amankwaa, Godfred; Amoako, Jones; Kaaratoore | Knowledge, attitude, and use of mHealth technology among students in Ghana: A university-based survey | 2019 | Digital Health | Ghana |

| | | | | | |
|---|---|---|---|---|---|
| 14 | Adatara P, Jonathan JWA., Baku EA, Atakro CA, Adedia DM | Factors Influencing Information and Communication Technology Knowledge and Use Among Nurse Managers in Selected Hospitals in the Volta Region of Ghana | 2019 | Digital Health | Ghana |
| 15 | Mitropoulos K, Patrinos GP., Cooper DN, Mitropoulou C, Agathos S, Reichardt JKV, Al-Maskari F, Chantratita W, Wonkam A, Dandara C, Katsila T, Lopez-Correa C, Ali BR | Genomic Medicine Without Borders: Which Strategies Should Developing Countries Employ to Invest in Precision Medicine? A New "Fast-Second Winner" Strategy | 2017 | Personalized Medicine | Low Resource Setting |
| 16 | Pagalday-Olivares P, Buendia R., BA, Adjordor-van de Beek J, Abudey S, Silberberg AR | Exploring the feasibility of eHealth solutions to decrease delays in maternal healthcare in remote communities of Ghana | 2017 | Digital Health | Ghana |
| 17 | Mensah, Isaac Kofi | Understanding the Drivers of Ghanaian Citizens' Adoption Intentions of Mobile Health Services | 2022 | Digital Health | Ghana |
| 18 | Ogoe HA, Douglas GP., Asamani JA, Hochheiser H | Assessing Ghana's eHealth workforce: | 2018 | Digital Health | Ghana |

| | | | | | |
|---|---|---|---|---|---|
| | | implications for planning and training | | | |
| 19 | Yusif S, Soar J., Hafeez-Baig A | An Exploratory Study of the Readiness of Public Healthcare Facilities in Developing Countries to Adopt Health Information Technology HITe-Health: the Case of Ghana | 2020 | Digital Health | Ghana |
| 20 | Amoakoh HB, Agyepong I., Klipstein-Grobusch K, Ansah EK, Grobbee DE, Yveoo L | How and why front-line health workers did not use a multifaceted mHealth intervention to support maternal and neonatal healthcare decision-making in Ghana | 2019 | Digital Health | Ghana |
| 21 | Peprah P, Akwasi AG., Abalo EM, Agyemang-Duah W, Budu HI, Appiah-Brempong E, Morgan AK | Lessening barriers to healthcare in rural Ghana: providers and users' perspectives on the role of mHealth technology. A qualitative exploration | 2020 | Digital Health | Ghana |
| 22 | Dzando G, Dordunu R., Akpeke H, Kumah A, Agada E, Lartey AA, Nortu J, Nutakor HS, Donyi AB | Telemedicine in Ghana: Insight into the past and present, a narrative review of literature | 2022 | Digital Health | Ghana |

| | | | | | |
|---|---|---|---|---|---|
| | | amidst the Coronavirus pandemic | | | |
| 23 | Sarfo, B, F; Treiber, F; Gebregziabher, M; Adamu, S; Patel, S; Nichols, M; Awuah, D; Sakyi, A; Adu-Darko, N; Singh, A; Tagge, R; Carolyn, J; Ovbiagele | PINGS Phone-Based Intervention Under Nurse Guidance After Stroke: interim Results of a Pilot Randomized Controlled Trial | 2018 | Digital Health | Ghana |
| 24 | D, Ben-Zeev | Mobile Health for Mental Health in West Africa: The Case for Ghana | 2018 | Digital Health | Ghana |
| 25 | Sarfo, B, FS; Treiber, F; Jenkins, C; Patel, S; Gebregziabher, M; Singh, A; Sarfo-Kantanka, O; Saulson, R; Appiah, L; Oparebea, E; Ovbiagele | Phone-based Intervention under Nurse Guidance after Stroke PINGS: study protocol for a randomized controlled trial | 2016 | Digital Health | Ghana |

| | | | | | |
|---|---|---|---|---|---|
| 26 | Sarfo, B, FS; Akpalu, A; Bockarie, A; Appiah, L; Nguah, SB; Ayisi-Boateng, NK; Adamu, S; Neizer, C; Arthur, A; Nyamekye, R; Agyenim-Boateng, K; Tagge, R; Adusei-Mensah, N; Ampofo, M; Laryea, R; Singh, A; Amuasi, JH; Ovbiagele | Phone-Based Intervention under Nurse Guidance after Stroke PINGS II Study: protocol for a Phase III Randomized Clinical Trial | 2021 | Digital Health | Ghana |
| 27 | L'Engle, S, KL; Green, K; Succop, SM; Laar, A; Wambugu | Scaled-Up Mobile Phone Intervention for HIV Care and Treatment: protocol for a Facility Randomized Controlled Trial | 2015 | Digital Health | Ghana |
| 28 | Barkman C, Weinehall L. | Policymakers and mHealth: roles and expectations, with observations from Ethiopia, Ghana and Sweden | 2017 | Digital Health | Ghana |
| 29 | Larnyo E, Appiah R., Dai B, Larnyo A, Nutakor JA, Ampon-Wireko S, Nkrumah ENK | Impact of Actual Use Behavior of Healthcare Wearable Devices on Quality of Life: A Cross-Sectional Survey of People with Dementia | 2022 | Digital Health | Ghana |

| | | | | | |
|---|---|---|---|---|---|
| | | and Their Caregivers in Ghana | | | |
| 30 | Effah Kaufmann E, Abdullah F., Tackie R, Pitt JB, Mba S, Akwetey B, Quaye D, Mills G, Nyame C, Bulley H, Glucksberg M, Ghomrawi H, Appeadu-Mensah W | Feasibility of Leveraging Consumer Wearable Devices with Data Platform Integration for Patient Vital Monitoring in Low-Resource Settings | 2024 | Digital Health | Ghana |
| 31 | Nichols M, Jenkins C., Sarfo FS, Singh A, Qanungo S, Treiber F, Ovbiagele B, Saulson R, Patel S | Assessing Mobile Health Capacity and Task Shifting Strategies to Improve Hypertension Among Ghanaian Stroke Survivors | 2017 | Digital Health | Ghana |
| 32 | Amoakoh, EK, HB; Klipstein-Grobusch, K; Agyepong, IA; Amoakoh-Coleman, M; Kayode, GA; Reitsma, JB; Grobbee, DE; Ansah | Can an mhealth clinical decision-making support system improve adherence to neonatal healthcare protocols in a low-resource setting? | 2020 | Digital Health | Ghana |

| | | | | | |
|---|---|---|---|---|---|
| 33 | Lee S, Kim SY., Lee YJ, Kim S, Choi W, Jeong Y, Rhim NJ, Seo I | Perceptions on Data Quality, Use, and Management Following the Adoption of Tablet-Based Electronic Health Records: Results from a Pre-Post Survey with District Health Officers in Ghana | 2022 | Digital Health | Ghana |
| 34 | Asgary R, Ogedegbe O., Adongo PB, Nwameme A, Cole HV, Maya E, Liu M, Yeates K, Adanu R | mHealth to Train Community Health Nurses in Visual Inspection With Acetic Acid for Cervical Cancer Screening in Ghana | 2016 | Digital Health | Ghana |
| 35 | Peprah, Adjei Gyimah, Prince; Abalo, Emmanuel Mawuli; Agyemang-Duah, Williams; Budu, Hayford Isaac; Appiah-Brempong, Emmanuel; Morgan, Anthony Kwame; Akwasi | Lessening barriers to healthcare in rural Ghana: providers and users' perspectives on the role of mHealth technology. A qualitative exploration | 2020 | Digital Health | Ghana |
| 36 | Mohammed, E, A; Acheampong, PR; Otupiri, E; Osei, FA; Larson-Reindorf, R; Owusu-Dabo | Mobile phone short message service SMS as a malaria control tool: a quasi-experimental study | 2019 | Digital Health | Ghana |

| | | | | | |
|---|---|---|---|---|---|
| 37 | Adadey SM, Wonkam A., Tingang Wonkam E, Twumasi Aboagye E, Quansah D, Asante-Poku A, Quaye O, Amedofu GK, Awandare GA | Enhancing Genetic Medicine: Rapid and Cost-Effective Molecular Diagnosis for a GJB2 Founder Mutation for Hearing Impairment in Ghana | 2020 | Personalized Medicine | Ghana |
| 38 | Rokicki S, Fink G. | Assessing the reach and effectiveness of mHealth: evidence from a reproductive health program for adolescent girls in Ghana | 2017 | Digital Health | Ghana |
| 39 | Ginsburg AS, Anderson R., Delarosa J, Brunette W, Levari S, Sundt M, Larson C, Tawiah Agyemang C, Newton S, Borriello G | mPneumonia: Development of an Innovative mHealth Application for Diagnosing and Treating Childhood Pneumonia and Other Childhood Illnesses in Low-Resource Settings | 2015 | Digital Health | Low Resource Setting |
| 40 | Mariwah S, Hampshire K., Machistey Abane A, Asiedu Owusu S, Kasim A, Robson E, Castelli M | Formalising 'informal' mHealth in Ghana: Opportunities and challenges for Universal Health Coverage UHC | 2022 | Digital Health | Ghana |

**Supplementary Table 3. List of Included Studies.** This table lists the final studies included in the scoping review, each identified by a unique key, title, and DOI, focusing on Digital health solutions, personalised medicine and N-of-1 trials in Ghana.

| Parameter | Value | Remarks |
| --- | --- | --- |
| Records identified from databases | 6282 | databases include: PubMed, IEEE Xplore, The Cochrane Library, Google Scholar, Government Websites - Ministry of Health, and Ghana Health Service, Directory of Open Access Journals, Other Organizations WHO, CDC, NGOs |
| Records identified from other sources | 13 | |
| Duplicate records removed | 2178 | |
| Records marked as ineligible by automation tools | 4057 | re categorization using keyword matching and machine learning algorithms in python |
| Records removed for other reasons | 1 | retracted articles |
| Records screened | 59 | |
| Records excluded | 0 | |
| Reports sought for retrieval | 59 | |
| Reports not retrieved | 10 | |
| Reports assessed for eligibility | 49 | |
| Reports excluded (reasons) | 9 | Does not meet Study objectives |
| Studies included in review | 40 | |
| Reports of included studies | 40 | |

**Supplementary Table 4. PRISMA Summary Table for Included Studies**
This table summarizes the document selection process for a scoping review according to the PRISMA guidelines, detailing each stage from identification to inclusion of studies.

| Database Regular Keyword Search (dataset) | | | |
|---|---|---|---|
| **Context** | **Digital Health** | **Personalised Medicine** | **Grand Total** |
| Ghana | 1604 | 614 | 2218 |
| Global | 1 | 1 | 2 |
| Low Resource Setting | 2497 | 1576 | 4073 |
| USA | 1 | 0 | 1 |
| **Grand Total** | **4103** | **2191** | **6294** |
| Raw Data extracted from Sources | | | |
| Python Normal Keyword Search (cleaned_dataset) | | | |
| **Context** | **Digital Health** | **Personalized Medicine** | **Grand Total** |
| Ghana | 81 | 2 | 366 |
| Low Resource Setting | 60 | 1 | 158 |
| **Grand Total** | **141** | **3** | **524** |
| Duplicates were removed at this stage | | | |
| Naïve Bayes (cleaned_dataset) | | | |
| **Context** | **Digital Health** | **Personalized Medicine** | **Grand Total** |
| Ghana | 79 | 3 | 82 |
| Low Resource Setting | 61 | 1 | 62 |
| **Grand Total** | **140** | **4** | **144** |
| Support Vector Machine (cleaned_dataset) | | | |
| Context | **Digital Health** | **Personalized Medicine** | **Grand Total** |
| Ghana | 82 | 2 | 84 |
| Low Resource Setting | 59 | 1 | 60 |
| **Grand Total** | **141** | **3** | **144** |
| Random Forest (cleaned_dataset) | | | |
| **Context** | **Digital Health** | **Personalized Medicine** | **Grand Total** |
| Ghana | 82 | 2 | 84 |
| Low Resource Setting | 59 | 1 | 60 |
| **Grand Total** | **141** | **3** | **144** |
| Ensemble Learning (cleaned_dataset) | | | |
| Context | **Digital Health** | **Personalized Medicine** | **Grand Total** |
| Ghana | 82 | 2 | 84 |
| Low Resource Setting | 59 | 1 | 60 |
| **Grand Total** | **141** | **3** | **144** |

| **Model Evaluation** | **Naïve Bayes** | **Support Vector Machine** | **Random Forest** | **Ensemble** |
|---|---|---|---|---|
| Context Model Accuracy | 0.97 | 0.97 | 0.97 | 0.97 |
| Theme Model Accuracy | 1.00 | 1.00 | 1.00 | 1.00 |

**Supplementary Table 5. Summary Results for Automated Document Screening**

This table describes the automated document screening process for our scoping review, using various machine learning models (Naïve Bayes, Support Vector Machine, Random Forest, Ensemble Learning) to refine search results by context and theme using the extracted meta data from databases and other sources.

### Supplementary Figures

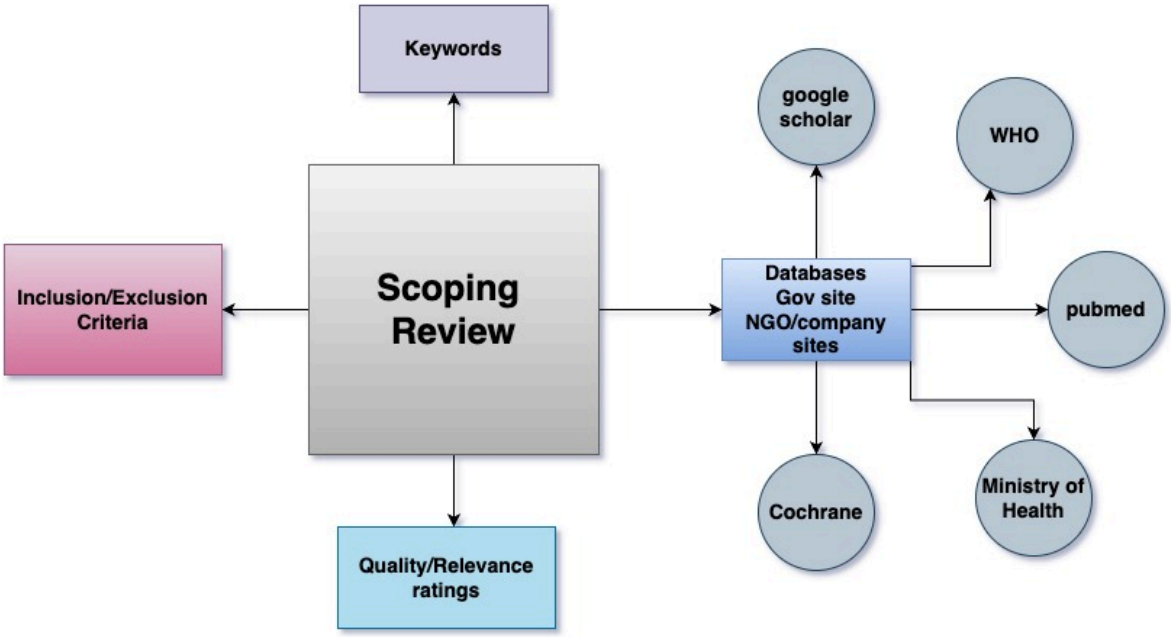

**Supplementary Figure 1. Review Approach**
This figure outlines the sources used for extracting papers and documents in the scoping review, including databases like PubMed, WHO, and Google Scholar.

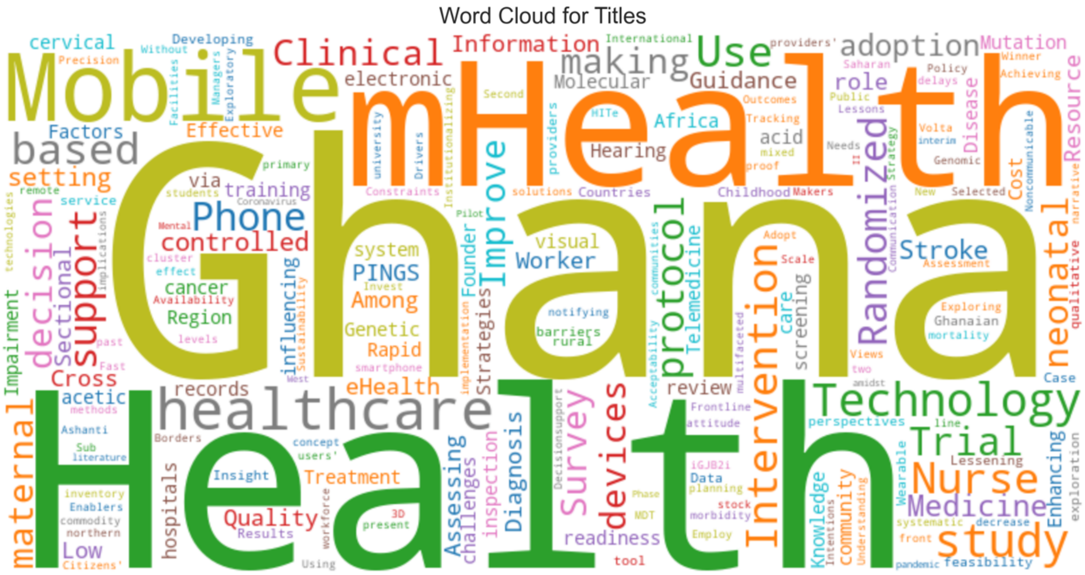

**Supplementary Figure 2. Keywords in paper titles**
This word cloud displays the key terms extracted from the titles of the selected papers.

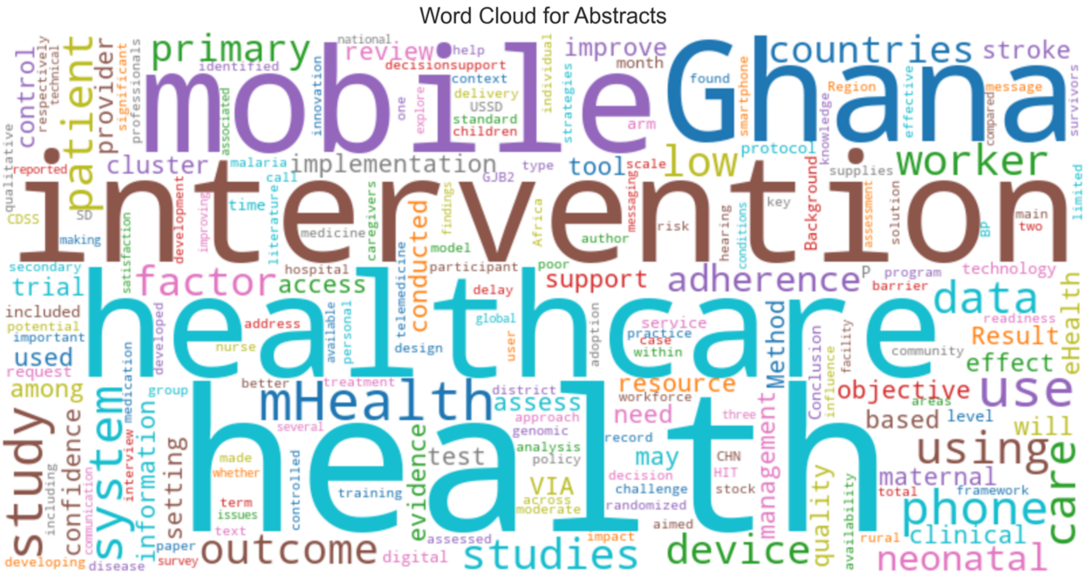

**Supplementary Figure 3. Keywords in paper Abstracts**
The word cloud displays the key terms extracted from the abstracts of the selected papers
